# Cannabis use prevalence before and early after partial legalisation in Germany

**DOI:** 10.64898/2026.03.18.26348683

**Authors:** Daniel Kotz, Jakob Manthey, Stephanie Klosterhalfen, Paula Steinhoff, Wolfgang Viechtbauer

## Abstract

**Background and Aims:** On April 1st, 2024, Germany implemented the Act on the Handling of Cannabis for Non-Medical Use (KCanG), allowing adults to cultivate and possess recreational cannabis. We assessed whether this policy shift was associated with a change in the prevalence of cannabis use in the general population and in daily or almost daily cannabis use.

**Design:** A series of 21 repeated cross-sectional surveys conducted between April/May 2022 and October/November 2025 (covering the period approximately two years before and one and a half years after the KCanG).

**Setting:** Population of Germany.

**Participants:** A total of 32,991 people aged 14-64 years, including 2,092 (6.3%) people who used cannabis in the past 12 months.

**Measurements:** Past 12-month cannabis use (at least once). In past 12-month users: daily or almost daily use. To test a potential change in prevalence following the KCanG, we used piecewise binomial logistic regression models using the exact date of each wave as the predictor variable, allowing for a change in the slope at the first full wave after implementation of the KCanG in April 2024, with a random effect for wave. We conducted this analysis for the total sample as well as stratified by gender (male vs. female), age (14-24 vs. 25-64 years), and for daily or almost daily cannabis use in the subgroup of people who used cannabis in the past 12 months. Sensitivity analyses used alternate intervention dates (in-time placebo tests).

**Findings:** The prevalence of cannabis use and the share of (almost) daily users among 12-month users remained largely stable before and after the law reform. None of the slope coefficients before the introduction of the KCanG were statistically significant (all p ≥ .08), and none of the coefficients for the change in the slope were statistically significant (all p ≥ .31). Results of sensitivity analyses confirmed the stable trends for both outcomes.

**Conclusions:** The legislation of cannabis introduced in Germany in April 2024 was not associated with a change in trends of 12-month cannabis use prevalence early (1.5 years) after implementation, and also not with a change in the proportion of heavy users among past-12-month users. We recommend continued close monitoring of trends using multiple data sources and over a longer post-implementation period, as the effects of the legislation may not have fully unfolded yet.

## BACKGROUND

In Germany, cannabis use prevalence has nearly doubled between 2012 and 2021, despite possession and supply being prohibited.[1] A similar trend has also been observed in other European countries.[2] In Germany in 2021, 12-month prevalence rates of usage were estimated at 8% in 12-to 17-year-olds, 25% in 18- to 25-year-olds, and 9% in 18- to 64-year-olds.[3, 4]

On April 1^st^, 2024, Germany implemented the Act on the Handling of Cannabis for Non-Medical Use (“Konsumcannabisgesetz”; KCanG).[5] The KCanG allows adults to possess and cultivate limited amounts for personal use and to form non-profit cannabis associations, while commercial sale remains prohibited. This policy shift, termed “partial legalisation” by the government, represents a departure from previous restrictive models but does not fully mirror the commercial legalisation seen in Canada or several US states. However, there has also been a shift in German medical cannabis policies in April 2024. Although medical cannabis has been legal since 2017, residents of Germany can now access cannabis flowers on prescription for almost any medical purpose via telemedicine. This has resulted in the largest commercial cannabis market in Europe, with around 200 tonnes of medical cannabis being available in 2025.[6]

Given these developments, there is a critical need to assess whether the prevalence of cannabis use in the German population has changed in response to the new law. This analysis is essential for understanding the public health implications of the reform and for informing future regulatory decisions. Our primary research question was: is the partial legalisation of cannabis, implemented in April 2024 in Germany, associated with a change in 12-month cannabis use prevalence in the general population and in selected subgroups (by age and gender)? Our secondary research question was: in the subgroup of people who used cannabis in the past 12 months, is the partial legalisation associated with a change in daily or almost daily cannabis use?

## METHODS

We used data from the German Study on Tobacco Use (DEBRA: “Deutsche Befragung zum Rauchverhalten”): an ongoing series of surveys on the use of nicotine, tobacco, and cannabis in the German population.[7] In a dual frame design, respondents are recruited either by random stratified sampling (approximately 40% of the sample) or quota sampling (https://osf.io/s2wxc). Data are collected through computer-assisted face-to-face interviews at home. The study received ethical approval from the Heinrich-Heine-University Düsseldorf (HHU 5386R) and was registered in the German Clinical Trials Register (registration numbers DRKS00011322, DRKS00017157, and DRKS00028054). A detailed plan for the current analyses was published prior to data analysis (https://osf.io/w92v5).

### Sample size

We use data from waves 36 (April/May 2022) to 56 (October/November 2025) of the DEBRA study; i.e., approximately two years before (waves 36-47; April 2022 to April 2024) and one and a half years after the KCanG was implemented (waves 48-56; May 2024 to November 2025). We restricted our analysis to people aged 14-64 years because, as of January 2025 (wave 52), we applied an upper age limit of 64 years during data collection.

### Measurements

We asked: “Have you ever used cannabis?” Responses “Yes, I have used cannabis daily or almost daily / at least once a week / at least once a month in the last 12 months / less than once a month” were defined as past 12-month cannabis use. Other measurements included years of age (14-24 vs. 25-64 years) and gender (male vs. female).

### Statistical analyses

We weighted data to approximate the sample to the German population in terms of age, gender, household size, level of education, and region (see [7] for details). We plotted the prevalence of past 12-month cannabis use (in %) for each wave along with pointwise 95% confidence intervals (CIs) (Clopper-Pearson method). Furthermore, we plotted the prevalence of daily or almost daily cannabis use in the subgroup of past 12-month users in the same way.

To smooth the raw data and to descriptively model trends in past 12-month cannabis use over the waves 36-56, we first used a binomial logistic regression model with a random effect for wave (to account for potential overdispersion). We utilised a restricted cubic spline term for wave (using the exact date at which 50% of the sample from that wave had been interviewed) with four knots at the following date positions: the midpoint between wave 40 and 41 (20 February 2023), the midpoint between wave 44 and 45 (06 October 2023), the midpoint between wave 47 and 48 (22 April 2024), and the midpoint between wave 50 and 51 (27 September 2024).

To address the primary research question – testing a potential change in prevalence implementation of the KCanG in April 2024 – we used a piecewise binomial logistic regression model using the exact date of each wave (date when 50% had been interviewed) as the predictor variable. The model tested for a change in the slope at wave 48 (i.e., the first full wave after April 2024 with field work starting 26 April 2024) and included a random effect for wave. We did not plan to model a level shift because such a policy effect on the population appears unlikely, and to avoid the risk of misclassifying noise in the data as a shift. We conducted in-time placebo tests as sensitivity analyses by moving the wave with a change in the slope to occur by ±2 waves (i.e., to wave 46, 47, 49, or 50).

Both trend analyses were conducted for the total sample as well as stratified by gender (male vs. female) and age (14-24 vs. 25-64 years). Furthermore, both trend analyses were also conducted for daily or almost daily cannabis use in the subgroup of people who used cannabis in the past 12 months (secondary research question). These subgroup analyses were not stratified by age and gender due to the small sample size of the subgroup.

All analyses were conducted in R (version 4.5.2). The data cannot be shared, but the statistical code is publicly available: https://osf.io/w92v5.

## RESULTS

In the total group of 32,991 respondents, 6.3% (n= 2,092) people reported past 12-month cannabis use. In this subgroup, 11.6% (n=242) reported daily or almost daily use. Out of the 2,092 people that reported past 12-month cannabis use, 1,491 were male, 601 female, 678 fell into the 14-24 age group, and 1,411 into the 25-64 age group.

Figure 1 describes the trend in the prevalence of past 12-month cannabis use. Overall, cannabis use prevalence remained largely stable over the study period (Figure 1a). In males (Figure 1b), the prevalence increased around the third quarter of the year 2023, then decreased, and then increased again from the middle of 2024 onwards. A similar but more pronounced pattern can be observed in 14- to 24-year-olds (Figure 1d). In the subgroup of people who used cannabis in the past 12 months, the prevalence of daily or almost daily cannabis use showed more fluctuations and overall, a slight decline (Figure 1f).

**Figure 1:**
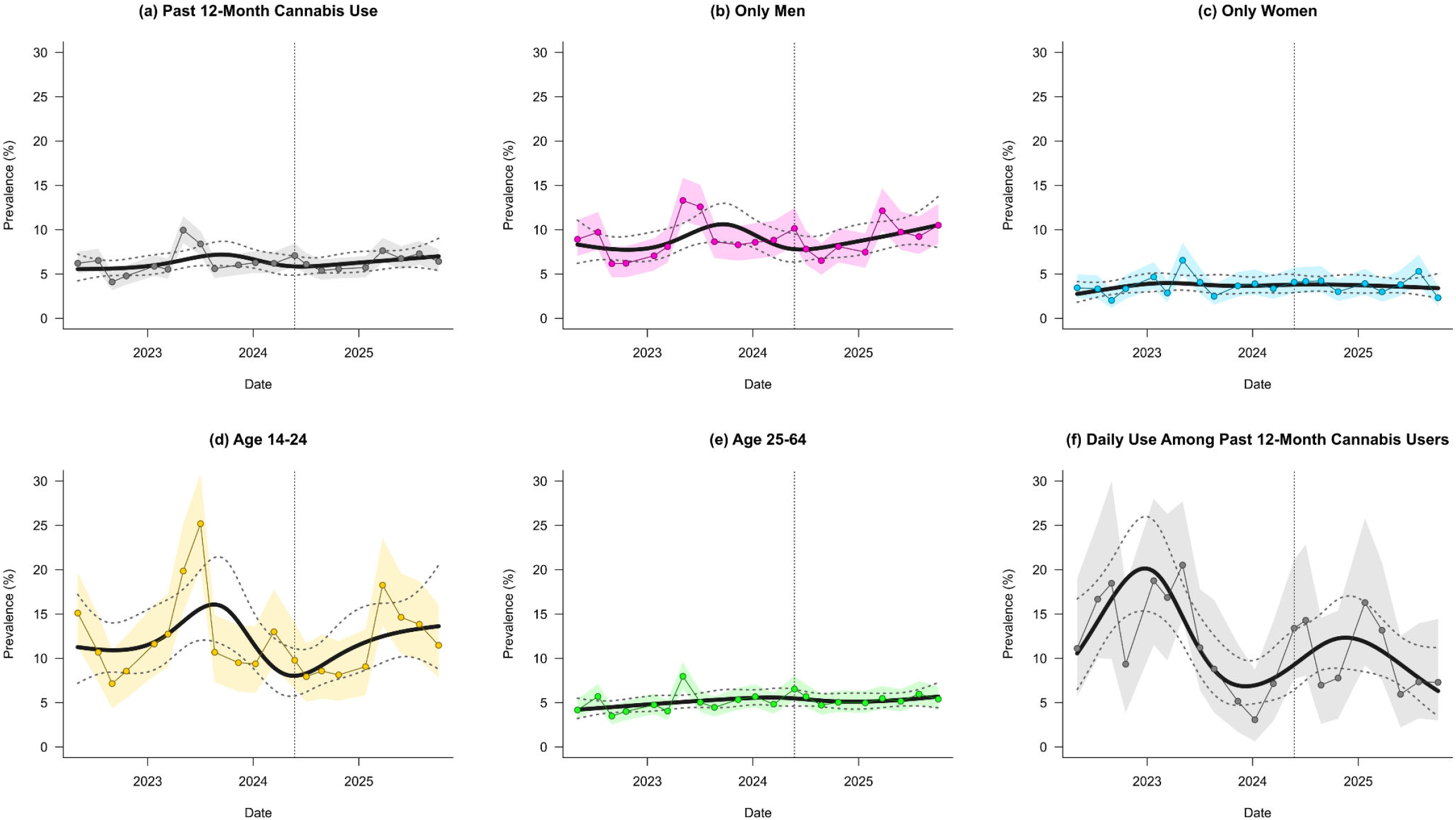
Trends in the prevalence of past 12-month cannabis use. The vertical dotted lines represent the date of implementation (April 1^st^, 2024) of the law regulating the handling and consumption of recreational cannabis.

Figure 2 shows the slopes of the piecewise binary logistic regression models in the total population (Figure 2a), and again stratified by gender (Figure 2b-c) and age (Figure 2d-e). The prevalence of daily or almost daily use in the subgroup of people who used cannabis in the past 12 months is shown in Figure 2f.

**Figure 2:**
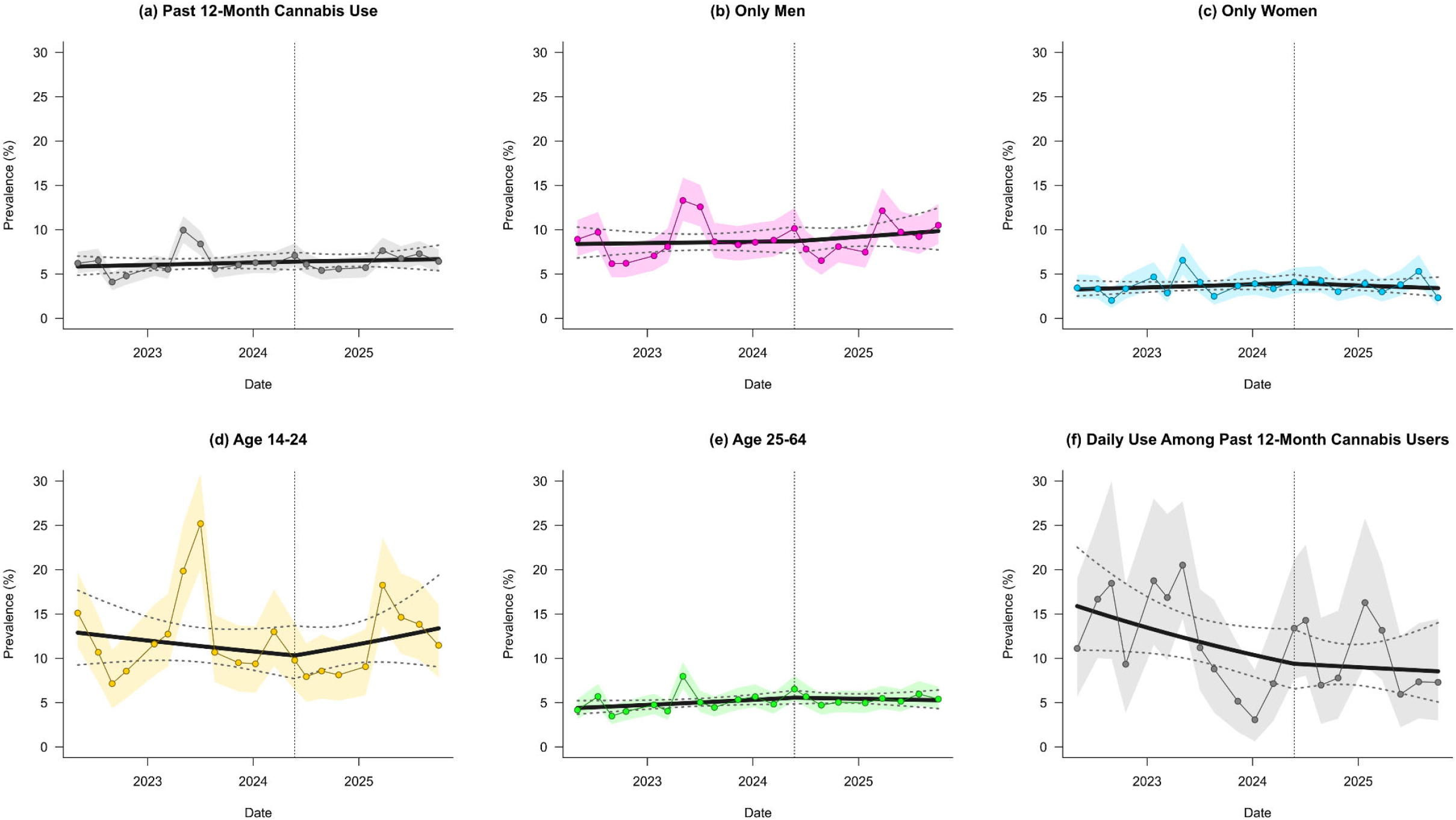
Piecewise binary logistic regression models of past 12-month cannabis use. The vertical dotted lines represent the date of implementation (April 1^st^, 2024) of the law regulating the handling and consumption of recreational cannabis.

Table 1 provides the regression coefficients for the main models. None of the slope coefficients before introduction of the KCanG were statistically significant (all p ≥ .08), and none of the coefficients for the change in the slope were statistically significant (all p ≥ .31). Moreover, none of the changes in the slope were significant in the sensitivity analyses (see Supplementary Table E1).

**Table 1:** Regression coefficients (with corresponding standard errors and p-values) for the piecewise binary logistic regression models. KCanG = Act on the Handling of Cannabis for Non-Medical Use (“Konsumcannabisgesetz”). Δ = change in the slope at wave 48 (i.e., after implementation of the KCanG in April 2024).

|  | All<br>(n=32,991) | Only<br>Men<br>(n=16,692) | Only<br>Women<br>(n=16,299) | Age<br>14-24<br>(n=5,599) | Age<br>25-64<br>(n=27,393) | Daily Use<br>(n=2,092) |
| --- | --- | --- | --- | --- | --- | --- |
| **Intercept** | -2.779<br>(0.0998)<br>p=0.00 | -2.391<br>(0.1155)<br>p=0.00 | -3.386<br>(0.1392)<br>p=0.00 | -1.911<br>(0.1901)<br>p=0.00 | -3.080<br>(0.0919)<br>p=0.00 | -1.667<br>(0.2204)<br>p=0.00 |
| **Slope until KCanG** | 0.013<br>(0.0203)<br>p=0.53 | 0.005<br>(0.0235)<br>p=0.82 | 0.027<br>(0.0281)<br>p=0.34 | -0.033<br>(0.0392)<br>p=0.39 | 0.033<br>(0.0185)<br>p=0.08 | -0.080<br>(0.0464)<br>p=0.08 |
| **ΔSlope after KCanG** | -0.004<br>(0.0474)<br>p=0.94 | 0.022<br>(0.0547)<br>p=0.68 | -0.059<br>(0.0660)<br>p=0.37 | 0.093<br>(0.0917)<br>p=0.31 | -0.044<br>(0.0427)<br>p=0.31 | 0.059<br>(0.1121)<br>p=0.60 |

## DISCUSSION

Using data from repeated cross-sectional population surveys before and early after partial legalisation of cannabis in Germany, we did not find a short-term change in trends of 12-month cannabis use prevalence in the general population and in selected subgroups (by age and gender). There was also no association with a change in daily or almost daily cannabis use in the subgroup of people who used cannabis in the past 12 months.

There are other studies on trends in the prevalence of cannabis use in Germany using representative surveys. A nationwide survey conducted every three years reported a steady increase in the 12-month prevalence in the age group 18-59 years from 5.2% in 2012 to 10.0% in 2021.[1] Similar trends were reported for both males and females.[1] The forecasted increase of use prevalence [1] was confirmed in more recent data from the same survey: among 18- to 64-year-olds, the 12-month prevalence was estimated at 8.8% in 2021 and at 9.8% in 2024 (after the reform); however, this additional increase was not statistically significant.[8] Furthermore, this additional increase was only found in males (from 10,7% in 2021 to 12,3% in 2024) but not in females (6,8% to 7,1%).[1, 8] Another general population survey with a focus on young people reported a steady increase in 12-month prevalence in the age group 18-25 years since 2008, again with similar trends for both males and females.[9] The study also identified use prevalence to have increased in males from 2023 to 2025 (26.9% to 31.6%), but not in females (19.4% to 18.8%).[9]

With regard to cannabis use patterns in the adult population, the proportion of people with heavy cannabis use (defined as daily or almost daily use or at least 200 times) among past 12-month users has remained fairly constant at around 10% since 1995, except for a moderately higher proportion in the year 2021 (15.7%).[1] More recent survey data reported no significant differences in the proportion of heavy use among adult users between 2021 (17.3%) and 2024 (16.2%).[8] Overall, these findings on trends in prevalence of cannabis use in the general population and in selected subgroups, and on heavy cannabis use are broadly consistent with our findings.

Research from international studies on the effects of recreational cannabis laws on the prevalence of cannabis use has produced mixed results. Some studies did and some did not find higher cannabis use among young adults.[10, 11] In the US, one study found increases in past-month and in frequent (20 days in past month) cannabis use in adults 26 years or older in states that legalised cannabis for non-medical purposes.[12] A more recent study found rising use among populations that have traditionally exhibited lower prevalence — such as older adults (≥60 years) and women — rather than among individuals who were already using cannabis.[13] In Canada, one study found that in the five years following legalisation, cannabis use frequency showed a modest increase, whereas cannabis misuse declined slightly.[14]

Our study has several limitations. Most importantly, our survey (like most surveys) does not represent marginalised groups of the population, and thus possible changes in use in certain populations (those most vulnerable) are probably not captured. Furthermore, the absolute prevalence of cannabis use in our study is likely to be underestimated due to the use of face-to-face interviews at home.[15] Moreover, bias with regard to the aim of the current study would arise if the legalisation of cannabis has altered the likelihood to disclose cannabis use among respondents. A key strength includes the collection of population data from personal interviews and a high frequency of data collection (21 data points in total, 12 before and 9 after the KCanG).

To conclude, the partial legislation of cannabis was not associated with a short-term change in 12-months cannabis use prevalence and also not with a change in the proportion of heavy users among past 12-month users. The increase in use prevalence observed since 2012 may continue with an attenuated slope. While the cannabis reform does not appear to have a noticeable short-term impact on use prevalence and patterns, we recommend continued close monitoring of trends using multiple data sources and over a longer post-implementation period. It appears likely that some policy effects — particularly regarding legal access through private and collective cultivation [6] — have not yet fully unfolded.

## Data Availability

All data produced in the present study are available upon reasonable request to the authors

https://osf.io/w92v5

---

**Supplementary Table E1:** Sensitivity analyses of the piecewise logistic regression models: coefficients for the change in the slope (with corresponding standard errors and p-values) after KCanG when shifting the intervention wave to waves 46-50.

|  | Wave 46 | Wave 47 | Wave 48 (main) | Wave 49 | Wave 50 |
| --- | --- | --- | --- | --- | --- |
| **All** | -0.019<br>(0.045)<br>p=0.677 | -0.011<br>(0.046)<br>p=0.804 | -0.004<br>(0.047)<br>p=0.938 | 0.004<br>(0.049)<br>p=0.936 | 0.015<br>(0.053)<br>p=0.781 |
| **Only Men** | -0.001<br>(0.053)<br>p=0.989 | 0.010<br>(0.053)<br>p=0.857 | 0.022<br>(0.055)<br>p=0.685 | 0.035<br>(0.057)<br>p=0.540 | 0.052<br>(0.061)<br>p=0.400 |
| **Only Women** | -0.057<br>(0.063)<br>p=0.367 | -0.055<br>(0.064)<br>p=0.390 | -0.059<br>(0.066)<br>p=0.371 | -0.061<br>(0.069)<br>p=0.374 | -0.063<br>(0.075)<br>p=0.402 |
| **Age 14-24** | 0.041<br>(0.089)<br>p=0.642 | 0.061<br>(0.089)<br>p=0.498 | 0.093<br>(0.092)<br>p=0.312 | 0.112<br>(0.095)<br>p=0.239 | 0.133<br>(0.102)<br>p=0.194 |
| **Age 25-64** | -0.046<br>(0.041)<br>p=0.261 | -0.043<br>(0.041)<br>p=0.303 | -0.044<br>(0.043)<br>p=0.305 | -0.040<br>(0.045)<br>p=0.371 | -0.033<br>(0.049)<br>p=0.506 |
| **Daily Use** | 0.118<br>(0.107)<br>p=0.270 | 0.097<br>(0.108)<br>p=0.368 | 0.059<br>(0.112)<br>p=0.597 | 0.046<br>(0.117)<br>p=0.693 | 0.045<br>(0.129)<br>p=0.726 |

KCanG = Act on the Handling of Cannabis for Non-Medical Use (“Konsumcannabisgesetz”). The coefficient represents the change in the time trend in prevalence after the assumed intervention wave compared with the pre-intervention trend. Models correspond to the main analysis but with the intervention wave shifted by $\pm 2$ waves relative to the main specification (wave 48).

